# On the Atrial Fibrillation Detection Performance of ECG-DualNet

**DOI:** 10.1101/2023.06.01.23290822

**Authors:** Christoph Reich, Maurice Rohr, Tim Kircher, Christoph Hoog Antink

**Affiliations:** Technische Universität Darmstadt; KIS*MED – AI Systems in Medicine, Technische Universität Darmstadt

## Abstract

Atrial Fibrillation (AF) is a severe cardiac arrhythmia affecting a significant amount of the human population [1]. Quick diagnosis and treatment are critical in reducing the risk of severe sequelae such as stroke or heart failure. Rohr *et al*. [2] recently proposed ECG-DualNet, a neural network for accurate AF detection in single-lead electrocardiogram (ECG) data. This short paper reports additional empirical results of ECG-DualNet to gain new insights on AF detection in single-lead ECG data with deep neural networks. We systematically analyze which ingredients of ECG-DualNet are crucial for achieving competitive AF detection results. We also scale the ECG-DualNet architecture to 130M parameters and perform large-scale supervised pre-training, providing additional empirical results. Finally, we provide recommendations for future research toward accurate and robust AF detection.

## I. ECG-DualNet Results

ECG-DualNet [2] is a two-branch network for AF detection in single-lead ECG data. The first branch (signal encoder) encodes the normalized time domain ECG signal. The second branch (spectrogram encoder) encodes the spectrogram of the ECG signal. ECG-DualNet utilizes an LSTM as the signal encoder and a ResNet as the spectrogram encoder. ECG-DualNet++ replaces the LSTM with a Transformer encoder and the ResNet convolutions with Axial-Attentions.

We utilize the PhysioNet dataset [3] to showcase the performance of ECG-DualNet(++). We split the publicly available data into 7000 training and 1528 validation samples. For the large-scale supervised pre-training we use the Icentia11K dataset [4], including *∼*550k single-lead ECG signals (*∼*500k training, *∼*50k validation). We perform 20 pre-training epochs on Icentia11K before fine-tuning on PhysioNet. Please refer to the original paper [2] for more architectural and training details.

Table I presents classification results and Floating Point Operations (FLOP) for different ECG-DualNet(++) variants. While being highly overparameterized ECG-DualNet++ 130M does not overfit and achieves a strong validation accuracy (0.8534), following the double descent phenomenon [5].

**TABLE I.** Classification (four classes) results on PhysioNet dataset.

| Model | ACC ↑ | F1 ↑ | FLOP | # Parameters |
| --- | --- | --- | --- | --- |
| ECG-DualNet S | 0.8527 | 0.8049 | 0.31G | 1.8 M |
| ECG-DualNet M | 0.8560 | 0.7938 | 1.04G | 4.3 M |
| ECG-DualNet L | 0.8514 | 0.8038 | 3.19G | 6.2 M |
| ECG-DualNet XL | **0.8612** | **0.8164** | 12.50G | 20.7 M |
| ECG-DualNet++ S | 0.8174 | 0.7291 | 0.72G | 1.8 M |
| ECG-DualNet++ M | 0.8259 | 0.7730 | 1.24G | 2.6 M |
| ECG-DualNet++ L | 0.8449 | 0.7859 | 2.94G | 3.7 M |
| ECG-DualNet++ XL | 0.8593 | 0.8051 | 9.77G | 8.2 M |
| ECG-DualNet++ 130M | 0.8534 | 0.7963 | 295.04G | 128.0 M |

While the signal encoder improves AF detection performance, interestingly, the most crucial part of the ECG-DualNet architecture is the spectrogram encoder (*c*.*f*. Table II). While data augmentation & dropout helps generalization we observe no severe overfitting when omitted.

**TABLE II.**
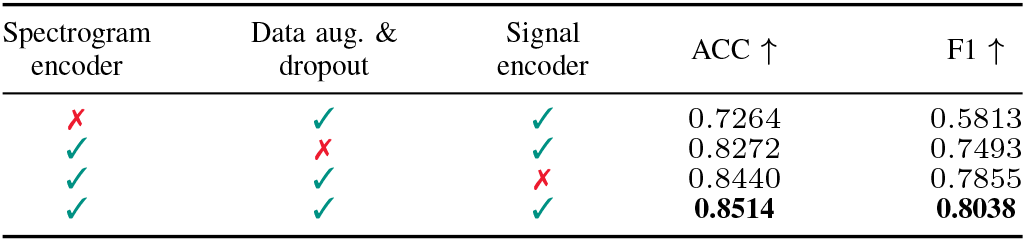
Ablation study on PhysioNet dataset w/ ECG-DualNet L.

| Spectrogram encoder | Data aug. & dropout | Signal encoder | ACC ↑ | F1 ↑ |
| --- | --- | --- | --- | --- |
| ✗ | ✓ | ✓ | 0.7264 | 0.5813 |
| ✓ | ✗ | ✓ | 0.8272 | 0.7493 |
| ✓ | ✓ | ✗ | 0.8440 | 0.7855 |
| ✓ | ✓ | ✓ | **0.8514** | **0.8038** |

Table III shows that large-scale supervised pre-training has no impact on the downstream performance on the PhysioNet dataset (*c*.*f*. Table I). We suspect a large domain shift between PhysioNet and Icentia11K caused by different ECG devices leading to this result. Future work might consider analyzing this behavior in more depth.

**TABLE III.** Results on PhysioNet dataset w/ supervised pre-training.

| Model | ACC ↑ | F1 ↑ |
| --- | --- | --- |
| ECG-DualNet XL | 0.8468 (↓ 0.0144) | 0.8014 (↓ 0.0150) |
| ECG-DualNet++ XL | 0.8527 (↓ 0.0066) | 0.7965 (↓ 0.0086) |

## II. CONCLUSION

We demonstrated, ECG-DualNet’s spectrogram encoder mostly contributes to a strong AF detection performance. Indicating that representing ECG data in the frequency domain is, in particular, suitable for AF detection with deep networks. Unifying the frequency and time domain within the network’s architecture might pose a possible direction for future research toward more efficient and robust AF detection.

## Data Availability

Both datasets used in the paper (2017 PhysioNet/CinC Challenge dataset & Icentia11k dataset) are available online at: https://physionet.org

